# Severe obesity is associated with higher in-hospital mortality in a cohort of patients with COVID-19 in the Bronx, New York

**DOI:** 10.1101/2020.05.05.20091983

**Authors:** Leonidas Palaiodimos, Damianos G. Kokkinidis, Weijia Li, Dimitrios Karamanis, Jennifer Ognibene, Shitij Arora, William N. Southern, Christos S. Mantzoros

## Abstract

**Background & Aims:** New York is the current epicenter of Coronavirus disease 2019 (COVID-19) pandemic. The underrepresented minorities, where the prevalence of obesity is higher, appear to be affected disproportionally. Our objectives were to assess the characteristics and early outcomes of patients hospitalized with COVID-19 in the Bronx and investigate whether obesity is associated with worse outcomes.

**Methods:** This retrospective study included the first 200 patients admitted to a tertiary medical center with COVID-19. The electronic medical records were reviewed at least three weeks after admission. The primary endpoint was in-hospital mortality.

**Results:** 200 patients were included (female sex: 102, African American: 102). The median BMI was 30 kg/m2. The median age was 64 years. Hypertension (76%), hyperlipemia (46.2%), and diabetes (39.5%) were the three most common comorbidities. Fever (86%), cough (76.5%), and dyspnea (68%) were the three most common symptoms. 24% died during hospitalization (BMI <25 kg/m2: 31.6%, BMI 25-34 kg/m2: 17.2%, BMI≥35 kg/m2: 34.8%, p= 0.03). The multivariate analysis for mortality, demonstrates that BMI≥35 kg/m2 (OR: 3.78; 95% CI: 1.45 - 9.83; p=0.006), male sex (OR: 2.74; 95% CI: 1.25 - 5.98; p=0.011) and increasing age (OR: 1.73; 95% CI: 1.13 - 2.63; p=0.011) were independently associated with higher in hospital mortality. Similar results were obtained for the outcomes of increasing oxygen requirement and intubation.

**Conclusions:** In this cohort of hospitalized patients with COVID-19 in a minority-predominant population, severe obesity, increasing age, and male sex were associated with higher in-hospital mortality and in general worse in-hospital outcomes.

## Introduction

Coronavirus disease 2019 (COVID-19), which is caused by severe acute respiratory syndrome coronavirus 2 (SARS-CoV-2), has evolved to a global pandemic with more than two million confirmed cases and about 200 thousand deaths so far [1]. The first cases in the United States (U.S.) were reported on January 19th, 2020 in the Washington state [2]. Since then, about one million confirmed cases and sixty thousand deaths have been reported [3]. New York City (NYC) is the current epicenter of the pandemic with about 160 thousand confirmed cases and more than twelve thousand deaths to date [4]. Early reports from Asia and Europe have identified older age, male sex, and chronic medical conditions, such as diabetes, hypertension, obesity, coronary artery disease, and heart failure, as risk factors associated with worse outcomes [5-7]. However, little is known about the risk factors and the characteristics of the disease in the United States population and particularly in underrepresented minorities, who appear to be affected disproportionally by COVID-19 [4,7]. The age-adjusted death rate per 100,000 people in NYC is more than double for African Americans compared to Whites (127.1 vs. 63.5) [4]. The higher prevalence of medical conditions that are considered risk factors for severe COVID-19 among African Americans and the higher risk for exposure to SARS-CoV-2 due to living and working conditions seem to be plausible explanations for the observed disproportionate differences in outcomes [7]. The Bronx, which is the most diverse area in the United States as per the 2010 U.S. Census, ranks last among all 62 counties of New York state in health outcomes, quality of life and important health and socioeconomic factors according to County Health Rankings and Roadmaps [8]. In addition, the Bronx has the highest obesity rates among all NYC boroughs and stands remarkably higher than the national average [9,10].

Our primary objective with this analysis was to investigate whether obesity is associated with worse in-hospital outcomes. Our secondary objective was to assess and present the clinical characteristics and early outcomes of the first 200 patients, who were diagnosed with COVID-19 and admitted to a large tertiary academic center and to define whether Diabetes, hypertension or other obesity comorbidities are independently associated with adverse outcomes.

## Materials and Methods

### Study design and patient population

This retrospective cohort study was conducted at the Montefiore Medical Center, a tertiary academic institution in the Bronx, New York. The first 200 patients who presented to the emergency room (ER) and were admitted to the inpatient medicine service or the intensive care unit (ICU) with laboratory-confirmed COVID-19 were included. We excluded patients who met one of the following exclusion criteria: i) discharge home directly from the ER, ii) transfer to our center after having received care in other institutions and iii) admission for non-COVID-19 related reasons or non-medical reasons (e.g. patients admitted because of a fracture, clinically stable patients residing in group homes unable to self-isolate). The 200 included patients were followed for three weeks after their admission to the hospital (admission of 1^st^ patient: March 9, 2020; admission of the 200th patient: March 22, 2020; completion of 3-week follow-up: April 12, 2020).

The study was approved by the institutional review board (IRB) of the Albert Einstein College of Medicine with a waiver of the inform consent (IRB number 2020-11296).

### Data Extraction

Two researchers (LP, WL) reviewed the electronic medical records (EMR) independently in a predefined data extraction sheet which was created for the purpose of this study. The documentation of the index admission from emergency medicine providers, inpatient providers, consultants, nurses, therapists, and social workers, the laboratory, and imaging data were reviewed. Post-discharge notes (e.g., tele-medicine follow-up visits, nursing outreach) were also reviewed when available. Documentation from past visits and the search engine of the EMR were also utilized.

The extracted data included baseline demographic information [(age, gender, race/ethnicity, residence status (community or skilled nursing facility/SNF), and zip code], clinical characteristics [body mass index (BMI), history of smoking, alcohol, intravenous drug use, hypertension, diabetes, hyperlipidemia, coronary artery disease (CAD), heart failure, cerebrovascular disease, chronic obstructive pulmonary disease (COPD), asthma, active malignancy, chronic kidney disease (CKD) or end-stage renal disease (ESRD), liver cirrhosis, and human immunodeficiency virus infection (HIV) or acquired immunodeficiency syndrome (AIDS)], pertinent home medications (immunosuppressive agents, ace-inhibitors, angiotensin II receptor blockers), symptomatology since disease onset and on presentation (fever, headache, malaise, myalgia, rhinorrhea, nasal congestion, sore throat, chest pain, dyspnea, cough, sputum production, nausea/vomiting, diarrhea), vital signs on presentation (oxygen saturation on room air, heart rate, presence of fever), level of oxygen requirement in the ER, laboratory data on the first hospital day [white cell count, lymphocyte count, hemoglobin, platelet count, creatinine, aspartate transaminase (AST), alanine transaminase (ALT), troponin T, creatine kinase (CPK), lactate dehydrogenase (LDH), Ferritin, d-dimers, C-reactive protein (CRP), procalcitonin, and hemoglobin A1c for diabetics], initial imaging findings (chest x-ray and/or chest computed tomography), oxygen requirements during hospital stay, acute respiratory distress syndrome (ARDS), intubation, number of days from presentation to intubation, ICU admission, acute kidney injury (AKI) or need for initiation of renal replacement therapy, length of stay, death, and hospital discharge.

The data were processed and analyzed without any personal identifiers to maintain patient confidentiality as per Health Insurance Portability and Accountability Act (HIPAA).

### Outcomes and Statistical Analysis

Patients were classified in three groups based on the BMI: BMI<25 kg/m^2^, BMI 25-34 kg/m^2^, and BMI≥35 kg/m^2^ as per the most recent BMI assessment prior to or during the index admission. Severe obesity was defined as BMI≥35 kg/m^2^. Patients were also classified in four quartiles based on age: ≤50, 51-64, 65-73, and ≥74 years old. The primary endpoint was in-hospital mortality. Secondary endpoints included: increasing oxygen requirement during hospital stay and intubation. Deceased patients were excluded from the length of stay analysis. Continuous data are presented as median with interquartile range (IQR) and categorical data as absolute and relative frequencies. The ANOVA test was used to compare the continuous variables, while chi-square was used for discrete variables. Interaction analyses were performed as needed. A logistic regression model was used to identify baseline variables associated with in-hospital mortality, intubation and increasing oxygen requirements. BMI 25-34 kg/m^2^ was used as a reference in order to perform dichotomous comparisons with patients with severe obesity (BMI≥ 35 kg/m^2^). In order to build a multivariate model, we used a forward stepwise approach with the following method for each one of the studied outcomes; model 1: BMI and age, model 2: all the variables with significant univariate associations (p value ≤ 0.05), and model 3: the variables of model 2 in addition to clinically significant variables which did not show a significant univariate association. Results of logistic regression are given as the odds ratio (OR) with the 95% confidence interval (CI). The threshold of statistical significance was p≤0.05. All analyses were performed using STATA software (version 14·1; STATA Corporation, College Station, TX, USA).

## Results

In total, 200 patients admitted with COVID-19 were included in this analysis (female sex= 102, BMI<25 kg/m^2^=38, BMI 25-34 kg/m^2^=116, and BMI≥35 kg/m^2^=46). The median BMI was 30 (IQR 26-35) kg/m^2^. Most of our patients were either of African American race (51%) or of Hispanic ethnicity (34.5%). 23.5% were SNF residents. The median age of the whole cohort was 64 (50-73.5) years, with significant differences among the three groups [BMI<25 kg/m^2^: 73 (64-80) vs. BMI 25-34 kg/m^2^ 63 [48.5-71] vs. BMI≥35 kg/m^2^: 57.5 (45-67), p<0.001). 32.5% of our cohort was active or past smokers. Hypertension, hyperlipemia and coronary artery disease were prevalent in 76%, 46.2% and 16.5% of our patients, respectively. 17% had a history of heart failure while 27.5% had a history of asthma or COPD. 29% percent had a history of chronic kidney disease or ESRD. Diabetes was prevalent in 39.5% of our patients. The detailed baseline demographic and clinical characteristics are presented in **Table 1**.

**Table 1.** Baseline demographic and clinical characteristics

| Characteristic | All patients | BMI group |  |  |  | Age group |  |  |  |  |
| --- | --- | --- | --- | --- | --- | --- | --- | --- | --- | --- |
|  | N=200 | BMI<25<br>(N=38) | BMI 25-34<br>(N=116) | BMI≥35<br>(N=46) | p-value | ≤50<br>(N=51) | 51-64<br>(N=53) | 65-73<br>(N=46) | ≥74<br>(N=50) | p-value |
|  |  | (a) | (b) | (c) |  | (a) | (b) | (c) | (d) |  |
| **Male sex - no. (%)** | 98 (49.0) | 21 (55) | 58 (50) | 19 (41) | 0.420 | 29 (56.9) | 20 (37.7) | 29 (63.0) | 20 (40.0) | 0.027 |
| **Age - years** |  |  |  |  |  |  |  |  |  |  |
| Median (IQR) | 64 (50-73.5) | 73 (64-80)<sup>bc</sup> | 63 (48.5-71)<sup>a</sup> | 57.5 (45-67)<sup>a</sup> | <0.001 | 42 (35-46) | 58 (56-62) | 68 (66-70) | 78 (75-84) | <0.001 |
| Distribution - no. (%) |  |  |  |  |  |  |  |  |  |  |
| ≤50 | 51 (25.5) | 4 (10.5)<sup>bc</sup> | 32 (27.6)<sup>a</sup> | 15 (32.6)<sup>a</sup> | <0.001 | 51 (100) | 0 (0.0) | 0 (0.0) | 0 (0.0) |  |
| 51-64 | 53 (26.5) | 6 (15.8)<sup>bc</sup> | 30 (25.9)<sup>a</sup> | 17 (7.0)<sup>a</sup> |  | 0 (0.0) | 53 (100) | 0 (0.0) | 0 (0.0) |  |
| 65-73 | 46 (23.0) | 9 (23.7)<sup>bc</sup> | 28 (24.1)<sup>a</sup> | 9 (19.6)<sup>a</sup> |  | 0 (0.0) | 0 (0.0) | 46 (100) | 0 (0.0) |  |
| ≥74 | 50 (25.0) | 19 (50.0)<sup>bc</sup> | 26 (22.4)<sup>a</sup> | 5 (10.9)<sup>a</sup> |  | 0 (0.0) | 0 (0.0) | 0 (0.0) | 50 (0.0) |  |
| **Residence status - no. (%)** |  |  |  |  |  |  |  |  |  |  |
| SNF resident | 47 (23.5) | 13 (34.2) | 25 (21.6) | 9 (19.6) | 0.216 | 4 (7.8)<sup>d</sup> | 11 (20.8) | 12 (26.1) | 20 (40.0)<sup>a</sup> | 0.002 |
| Community-based | 153 (76.5) | 25 (65.8) | 91 (78.5) | 37 (80.4) |  | 47 (92.2)<sup>d</sup> | 42 (79.2) | 34 (73.9) | 30 (60.0)<sup>a</sup> |  |
| **Race/Ethnicity - no. (%)** |  |  |  |  |  |  |  |  |  |  |
| African American | 102 (51.0) | 21 (55.3) | 55 (47.4) | 26 (56.5) | 0.142 | 18 (35.3)<sup>bd</sup> | 29 (54.7)<sup>a</sup> | 22 (47.8) | 33 (66.0)<sup>a</sup> | 0.004 |
| Hispanic/Latino | 69 (34.5) | 8 (21.1) | 47 (40.5) | 14 (30.4) |  | 39 (56.9)<sup>bd</sup> | 15 (28.3)<sup>a</sup> | 16 (34.8) | 9 (18.0)<sup>a</sup> |  |
| Other | 29 (14.5) | 9 (23.7) | 14 (12.1) | 6 (13.0) |  | 4 (7.8)<sup>bd</sup> | 9 (20.0)<sup>a</sup> | 8 (17.4) | 8 (16.0)<sup>a</sup> |  |
| **BMI - kg/m<sup>2</sup>** |  |  |  |  |  |  |  |  |  |  |
| Median (IQR) | 30 (26-35) | 22 (20.7-24)<sup>bc</sup> | 29 (27-31)<sup>ac</sup> | 41 (37-46)<sup>ab</sup> | <0.001 | 31 (27-38)<sup>d</sup> | 32 (29-37)<sup>d</sup> | 29 (25-32) | 26 (23-30)<sup>ab</sup> | <0.001 |
| **Smoking - no./total no. (%)** |  |  |  |  |  |  |  |  |  |  |
| Never smoked | 135 (67.5) | 20 (52.6) | 80 (69.0) | 35 (76.1) | 0.064 | 41 (80.4) | 32 (60.4) | 26 (56.5) | 36 (72.0) | 0.044 |
| Former or current smoker | 65 (32.5) | 18 (47.4) | 36 (31.0) | 11 (23.9) |  | 10 (19.6) | 21 (39.6) | 20 (43.5) | 14 (28.0) |  |
| **Coexisting disorder - no. (%)** |  |  |  |  |  |  |  |  |  |  |
| Any | 158 (79.0) | 32 (84.2) | 88 (75.9) | 38 (82.61) | 0.434 | 29 (57)<sup>bcd</sup> | 44 (83.0)<sup>a</sup> | 41 (89)<sup>a</sup> | 44 (88.0)<sup>a</sup> | <0.001 |
| Hypertension | 152 (76.0) | 30 (79.0) | 89 (76.7) | 33 (71.7) | 0.715 | 25 (49.0)<sup>bcd</sup> | 41 (77.4)<sup>a</sup> | 40 (87.0)<sup>a</sup> | 46 (92.0)<sup>a</sup> | <0.001 |
| Diabetes | 79 (39.5) | 14 (36.8) | 41 (35.3) | 24 (52.2) | 0.133 | 14 (27.5) | 21 (39.6) | 24 (52.2) | 20 (40.0) | 0.102 |
| Hyperlipidemia | 92 (46.2) | 16 (43.2) | 55 (47.4) | 21 (45.7) | 0.903 | 15 (29.4)<sup>c</sup> | 23 (43.4) | 27 (60.0)<sup>a</sup> | 27 (50.0) | 0.014 |
| Coronary artery disease | 33 (16.5) | 8 (21.1) | 19 (16.4) | 6 (13.0) | 0.615 | 3 (5.9)<sup>d</sup> | 10 (18.9) | 7 (15.2) | 13 (26.0)<sup>a</sup> | 0.052 |
| Cerebrovascular disease | 22 (11.0) | 9 (23.7)<sup>bc</sup> | 11 (9.5)<sup>a</sup> | 2 (4.4)<sup>a</sup> | 0.014 | 1 (2.0)<sup>d</sup> | 3 (5.7)<sup>d</sup> | 6 (13.0) | 12 (24.0)<sup>ab</sup> | 0.002 |
| Heart failure | 34 (17.0) | 14 (36.8)<sup>bc</sup> | 12 (10.3)<sup>a</sup> | 8 (17.4)<sup>a</sup> | 0.001 | 4 (7.8)<sup>d</sup> | 8 (15.1) | 7 (15.2) | 15 (30.0)<sup>a</sup> | 0.026 |
| Asthma | 27 (13.5) | 5 (13.2) | 18 (15.5) | 4 (8.7) | 0.518 | 10 (19.6) | 4 (7.6) | 9 (19.6) | 4 (8.0) | 0.112 |
| COPD | 28 (14.0) | 7 (18.4) | 14 (12.1) | 7 (15.2) | 0.597 | 0 (0.0)<sup>d</sup> | 10 (18.9) | 7 (15.2)<sup>d</sup> | 11 (22.0)<sup>ac</sup> | 0.007 |
| Chronic renal disease | 58 (29.0) | 16 (42.1) | 28 (24.1) | 14 (30.4) | 0.103 | 7 (13.7)<sup>d</sup> | 15 (28.3) | 16 (34.8) | 20 (40.0)<sup>a</sup> | 0.024 |
| CKD III-V | 41 (20.5) | 9 (23.7) | 20 (17.2) | 12 (26.1) |  | 3 (5.88) | 9 (16.7) | 13 (28.3) | 16 (32.0) | 0.033 |
| ESRD | 17 (8.5) | 7 (18.4) | 8 (6.9) | 2 (4.4) |  | 4 (7.8) | 6 (11.3) | 3 (6.5) | 4 (8.0) |  |
| Active malignancy | 11 (5.5) | 1 (2.6) | 6 (5.2) | 4 (8.7) | 0.465 | 0 (0.0)<sup>d</sup> | 3 (5.7) | 2 (4.4) | 6 (12.0)<sup>a</sup> | 0.067 |
| Liver cirrhosis | 2 (1.0) | 0 (0.0) | 0 (0.0)<sup>c</sup> | 2 (4.4)<sup>b</sup> | 0.034 | 0 (0.0) | 1 (1.89) | 1 (2.2) | 0 (0.0) | 0.556 |
| HIV/AIDS | 5 (2.5) | 1 (2.6) | 3 (2.6) | 1 (2.2) | 0.987 | 1 (2.0) | 3 (5.7) | 1 (2.2) | 0 (0.0) | 0.316 |
| **ACEi or ARB use - no. (%)** |  |  |  |  |  |  |  |  |  |  |
| ACEi | 36 (18.0) | 8 (21.1) | 20 (17.2) | 8 (17.4) | 0.862 | 5 (9.8) | 12 (22.6) | 9 (19.6) | 10 (20.0) | 0.347 |
| ARB | 25 (12.6) | 5 (13.2) | 14 (12.1) | 6 (13.3) | 0.969 | 5 (9.8) | 4 (7.7) | 6 (13.0) | 10 (20.0) | 0.261 |
| None | 138 (69.0) | 13 (34.2) | 34 (29.1) | 15 (32.6) | 0.821 | 41 (80.4) | 36 (67.9) | 31 (67.4) | 30 (60.0) | 0.167 |
| **Immunosuppressive tx - no. (%)** | 17 (8.5) | 3 (7.9) | 13 (11.2) | 1 (2.2) | 0.176 | 3 (5.9) | 5 (9.4) | 5 (10.9) | 4 (8.0) | 0.836 |

Note: p-values refer to Chi-square test/ANOVA and the letters denote the columns where a statistically significant pairwise comparison exists (Bonferroni's method)
Abbreviations and symbols: BMI=body mass index, IQR=interquartile range, no.=number, SNF=skilled nursing facility, kg=kilogram, m=meter, CKD=chronic kidney disease, ESRD=end-stage renal disease, HIV=human immunodeficiency virus, AIDS=acquired immunodeficiency syndrome, ACEi= angiotensin-converting enzyme inhibitor, ARB=angiotensin receptor blocker, tx=treatment

Fever (86%), cough (76.5%), dyspnea (68%) and malaise (58%) were the four most common symptoms. The median SO_2_ on the first hospital day was 95% (IQR 89-97) without significant differences among groups. Symptoms and signs are presented in **Table 2**. All 200 patients received chest x-ray on presentation, with 55.5% of them having bilateral infiltrates and 27% having only unilateral findings. The laboratory and radiologic findings are presented in **Table 3**.

**Table 2.** Symptoms and signs on presentation

| Characteristic | All patients | BMI group |  |  |  | Age group |  |  |  |  |
| --- | --- | --- | --- | --- | --- | --- | --- | --- | --- | --- |
|  | N=200 | BMI<25<br>(N=38) | BMI 25-34<br>(N=116) | BMI≥35<br>(N=46) | p-value | ≤50<br>(N=51) | 51-64<br>(N=53) | 65-73<br>(N=46) | ≥74<br>(N=50) | p-value |
|  |  | (a) | (b) | (c) |  | (a) | (b) | (c) | (d) |  |
| **Symptoms - no. (%)** |  |  |  |  |  |  |  |  |  |  |
| Fever | 172 (86.0) | 29 (16.3) | 102 (87.9) | 41 (89.1) | 0.158 | 48 (94.1)<sup>d</sup> | 47 (88.7) | 40 (87.0) | 37 (74.0)<sup>a</sup> | 0.028 |
| Cough | 153 (76.5) | 29 (76.3) | 88 (75.9) | 36 (78.3) | 0.948 | 42 (82.4) | 38 (71.7) | 37 (80.4) | 36 (72.0) | 0.456 |
| Dyspnea | 136 (68.0) | 23 (60.5) | 76 (65.5) | 37 (80.4) | 0.102 | 37 (72.6) | 39 (73.6) | 30 (65.2) | 30 (60.0) | 0.411 |
| Malaise | 116 (58.0) | 22 (57.9) | 70 (60.3) | 24 (52.2) | 0.637 | 26 (51.0) | 34 (64.2) | 29 (63.0) | 27 (54.0) | 0.446 |
| Diarrhea | 66 (33.0) | 8 (21.0)<sup>c</sup> | 35 (30.2)<sup>c</sup> | 23 (50.0)<sup>ab</sup> | 0.012 | 23 (45.1) | 21 (39.6) | 10 (21.7) | 12 (24.0) | 0.031 |
| Myalgia | 61 (30.5) | 9 (23.7) | 37 (31.9) | 15 (32.6) | 0.596 | 22 (43.1) | 17 (32.1) | 13 (28.3) | 9 (18.0) | 0.053 |
| Sputum production | 46 (23.0) | 8 (21.5) | 26 (22.4) | 12 (26.1) | 0.839 | 12 (23.5) | 15 (28.3) | 11 (23.9) | 8 (16.0) | 0.521 |
| Headache | 40 (20.0) | 4 (10.53) | 26 (22.4) | 10 (21.7) | 0.267 | 12 (23.5) | 17 (32.1)<sup>d</sup> | 9 (19.6) | 2 (4.0)<sup>b</sup> | 0.004 |
| Nasal congestion or rhinorrhea | 37 (18.5) | 7 (18.4) | 22 (19.0) | 8 (17.4) | 0.973 | 14 (27.5) | 11 (20.8) | 7 (15.2) | 5 (10.0) | 0.132 |
| Nausea or vomiting | 35 (17.5) | 4 (10.5) | 22 (19.0) | 9 (19.6) | 0.452 | 11 (21.6) | 10 (18.9) | 5 (10.9) | 9 (18.0) | 0.559 |
| Sore throat | 20 (10.0) | 4 (10.3) | 12 (10.3) | 4 (8.7) | 0.945 | 6 (11.8) | 7 (13.2) | 3 (6.5) | 4 (8.0) | 0.654 |
| **Recorded fever on day 1 - no. (%)** | 126 (63.0) | 22 (57.9) | 75 (64.7) | 29 (63.0) | 0.755 | 34 (66.7) | 34 (64.2) | 31 (67.4) | 27 (54.0) | 0.486 |
| **Minimum SO<sub>2</sub> on room air on day 1** |  |  |  |  |  |  |  |  |  |  |
| Median (IQR) - % | 95 (89-97) | 95 (87-97) | 95 (93-97) | 93 (88-96) | 0.346 | 96 (94-97) | 94 (89.5-97) | 94 (87-97) | 95 (88-96) | 0.077 |
| Distribution - no. (%) |  |  |  |  |  |  |  |  |  |  |
| ≤80% | 14 (7.0) | 1 (2.6) | 8 (6.9) | 5 (10.9) | 0.003 | 1 (2.0) | 5 (9.4) | 6 (13.0) | 2 (4.0) | 0.407 |
| 81-87% | 25 (12.5) | 10 (26.3) | 10 (8.6) | 5 (10.9) |  | 4 (7.8) | 6 (11.3) | 7 (15.2) | 8 (16.0) |  |
| 88-91% | 20 (10.0) | 4 (10.5) | 6 (5.2) | 10 (21.7) |  | 4 (7.84) | 6 (11.3) | 2 (4.4) | 8 (16.0) |  |
| 92-96% | 86 (43.0) | 11 (29.0) | 58 (50.0) | 17 (37.0) |  | 25 (49.0) | 22 (41.5) | 18 (39.1) | 21 (42.0) |  |
| ≥97% | 55 (27.5) | 12 (31.6) | 34 (29.3) | 9 (19.6) |  | 17 (33.3) | 14 (26.4) | 13 (28.3) | 11 (22.0) |  |

Note: p-values refer to Chi-square test/ANOVA and the letters denote the columns where a statistically significant pairwise comparison exists (Bonferroni's method)
Abbreviations: BMI=body mass index, IQR=interquartile range, no.=number, SO<sub>2</sub>=oxygen saturation

**Table 3.** Laboratory and imaging findings on presentation

| Characteristic | All patients | BMI group |  |  |  | Age group |  |  |  |  |
| --- | --- | --- | --- | --- | --- | --- | --- | --- | --- | --- |
|  | N=200 | BMI<25<br>(N=38) | BMI 25-34<br>(N=116) | BMI≥35<br>(N=46) | p-value | ≤50<br>(N=51) | 51-64<br>(N=53) | 65-73<br>(N=46) | ≥74<br>(N=50) | p-value |
|  |  | (a) | (b) | (c) |  | (a) | (b) | (c) | (d) |  |
| White-cell count |  |  |  |  |  |  |  |  |  |  |
| Median (IQR) - per 10<sup>3</sup>/uL | 6.3 (4.7-7.8) | 6.3 (4-8.3) | 6.1 (4.7-7.6) | 6.8 (5.3-8) | 0.075 | 5.7 (4.4-7.5) | 6.8 (5.2-8.8) | 6.2 (4.7-8.3) | 6.3 (5-7.4) | 0.622 |
| Distribution |  |  |  |  |  |  |  |  |  |  |
| ≥10,000 / uL | 23 (11.5) | 5 (13.2) | 13 (11.2) | 5 (10.9) | 0.937 | 4 (7.8) | 10 (18.9) | 5 (10.9) | 4 (8.0) | 0.249 |
| ≤4000/uL | 35 (17.5) | 10 (26.3) | 20 (17.2) | 5 (10.9) | 0.178 | 9 (17.7) | 8 (15.1) | 7 (15.2) | 11 (22.0) | 0.782 |
| Lymphocyte count |  |  |  |  |  |  |  |  |  |  |
| Median (IQR) - per 10<sup>3</sup>/uL | 0.9 (0.7-1.3) | 0.8 (0.6-1.1) | 0.9 (0.6-1.3) | 1.1 (0.8-1.5) | 0.467 | 0.9 (0.8-1.3) | 1 (0.7-1.3) | 0.9 (0.6-1.4) | 0.9 (0.6-1.3) | 0.923 |
| ≤1000/uL | 123 (61.5) | 28 (73.7)<sup>c</sup> | 73 (62.9) | 22 (47.8)<sup>a</sup> | 0.047 | 33 (64.7) | 28 (52.8) | 30 (65.2) | 32 (64.0) | 0.512 |
| Hemoglobin - Median (IQR) - g/dL | 12.7 (11.1-14.2) | 12 (10.3-13.8) | 13 (11.5-14.3) | 13 (10.8-14.1) | 0.667 | 14 (12.1-15.3)<sup>d</sup> | 12.7 (11-13.9) | 12.9 (11.4-14.1) | 12.2 (11-13.1)<sup>a</sup> | 0.029 |
| Platelets - Median (IQR) - per 10<sup>3</sup>/uL | 194 (149-240) | 178 (134-271) | 194 (154-240) | 208 (160-231) | 0.044 | 197 (167-231) | 200 (151-254) | 185 (144-232) | 177 (132-240) | 0.669 |
| Creatinine-Median (IQR) - mg/dL | 1 (0.8-1.7) | 1.15 (0.8-2.4) | 1 (0.8-1.5) | 1.1 (0.8-1.7) | 0.082 | 0.9 (0.7-1.2) | 0.9 (0.8-1.7) | 1.1 (0.8-1.9) | 1.2 (0.9-1.8) | 0.735 |
| AST≥50 U/L | 72 (36.0) | 15 (39.5) | 40 (34.5) | 17 (37.0) | 0.847 | 18 (35.3) | 23 (43.4) | 12 (26.1) | 19 (38.0) | 0.345 |
| ALT≥50 U/L | 36 (18.0) | 6 (15.8) | 22 (19.0) | 8 (17.4) | 0.900 | 11 (21.6) | 14 (26.4) | 6 (13.0) | 5 (10.0) | 0.116 |
| CK≥200 U/L | 104 (52.0) | 18 (47.4) | 56 (48.3) | 30 (65.2) | 0.123 | 21 (41.2) | 31 (58.5) | 25 (54.4) | 27 (54.0) | 0.325 |
| Troponin T≥0.1 ng/mL | 56 (28.0) | 14 (36.8) | 32 (27.6) | 10 (21.7) | 0.305 | 17 (33.3) | 16 (30.2) | 10 (21.7) | 13 (26.0) | 0.606 |
| LDH≥240 U/L | 172 (86.0) | 33 (86.8) | 97 (83.6) | 42 (91.3) | 0.440 | 47 (92.2) | 46 (86.8) | 38 (82.6) | 41 (82.0) | 0.434 |
| C-reactive protein |  |  |  |  |  |  |  |  |  |  |
| Median (IQR) - mg/dL | 8.35 (4.4-15) | 11.6 (5.5-24.6) | 7.75 (4.35-13.9) | 9.45 (4.5-14.95) | 0.025 | 8.1 (4.4-14.5) | 6.5 (4.4-12.2) | 11.2 (4.6-18.8) | 8 (3.5-15.6) | 0.362 |
| ≥5 mg/dL | 81/122<br>(66.4) | 17/22<br>(77.3) | 48/76<br>(63.2) | 16/24<br>(66.7) | 0.467 | 23/34<br>(67.6) | 21/33<br>(63.6) | 18/25<br>(72.0) | 19/30<br>(63.3) | 0.893 |
| ≥10 mg/dL | 55/122<br>(45.1) | 12/22<br>(54.6) | 31/76<br>(40.8) | 12/24<br>(50.0) | 0.450 | 16/34<br>(47.1) | 11/33<br>(33.3) | 14/25<br>(56.0) | 14/30<br>(46.7) | 0.372 |
| ≥15 mg/dL | 31/122<br>(25.4) | 9/22 (40.9) | 16/76<br>(21.1) | 6/24 (25.0) | 0.169 | 8/34 (23.5) | 6/33 (18.2) | 8/33 (32.0) | 9/33 (30.0) | 0.598 |
| D-dimer≥1 ug/mL | 38/64<br>(59.4) | 11/13<br>(84.6) | 19/37<br>(51.4) | 8/14 (57.1) | 0.108 | 9/21 (42.3) | 11/16<br>(68.75) | 8/13 (61.6) | 10/14<br>(71.4) | 0.281 |
| D-dimer≥3 ug/mL | 13/64<br>(20.3) | 2/13 (15.4) | 10/37<br>(27.0) | 1/14 (7.1) | 0.256 | 2/21 (9.5) | 5/16 (31.3) | 2/13 (15.4) | 4/14 (28.6) | 0.324 |
| Ferritin≥500 ng/mL | 14/22<br>(63.6) | 3/3<br>(100.0)<sup>c</sup> | 10/12<br>(83.3)<sup>c</sup> | 1/7<br>(14.3)<sup>ab</sup> | 0.004 | 4/5 (80.0) | 3/7 (42.9) | 6/8 (75.0) | 1/2 (50.0) | 0.477 |
| Ferritin>270 ng/mL | 20/22<br>(90.9) | 3/3 (100.0) | 12/12<br>(100.0) | 5/7 (71.4) | 0.095 | 4/5 (80.0) | 6/7 (85.7) | 8/8 (100) | 2/2 (100) | 0.583 |
| Procalcitonin |  |  |  |  |  |  |  |  |  |  |
| Median (IQR) - ng/mL | 0.1 (0.1-0.4) | 0.1 (0.1-0.8) | 0.1 (0.1-0.3) | 0.1 (0.1-0.6) | 0.544 | 0.1 (0.1-0.6) | 0.1 (0.1-0.6) | 0.2 (0.1-0.3) | 0.1 (0.1-0.3) | 0.346 |
| >0.1 ng/mL | 42/98<br>(42.9) | 9/19 (47.4) | 23/58<br>(40.0) | 10/21<br>(47.6) | 0.743 | 6/17 (35.3) | 13/33<br>(39.4) | 13/26<br>(50.0) | 10/22<br>(45.5) | 0.762 |
| ≥0.25 ng/mL | 31/98<br>(31.6) | 6/19 (31.6) | 17/58<br>(29.3) | 8/21 (38.1) | 0.759 | 5/17 (29.4) | 12/33<br>(36.4) | 7/26 (26.9) | 7/22 (31.8) | 0.886 |
| ≥0.5 ng/mL | 24/98<br>(24.5) | 6/19 (31.6) | 11/58<br>(19.0) | 7/21 (33.3) | 0.307 | 5/17 (29.4) | 9/33 (27.3) | 5/26 (19.2) | 5/22 (22.7) | 0.853 |
| ≥1 ng/mL | 18/98<br>(18.4) | 4/19 (21.1) | 10/58<br>(17.2) | 4/21 (19.1) | 0.929 | 4/17 (23.5) | 8/33 (24.2) | 1/26 (3.9) | 5/22 (22.7) | 0.172 |
| **Imaging on admission** |  |  |  |  |  |  |  |  |  |  |
| Chest radiography |  |  |  |  |  |  |  |  |  |  |
| No infiltrates | 35 (17.5) | 6 (15.8) | 22 (19.0) | 7 (15.2) | 0.085 | 9 (17.7) | 10 (18.9) | 9 (19.6) | 7 (14.0) | 0.300 |
| Unilateral infiltrates | 54 (27.0) | 17 (44.7) | 27 (23.3) | 10 (21.7) |  | 13 (25.5) | 9 (16.9) | 12 (26.1) | 20 (40.0) |  |
| Bilateral infiltrates | 111 (55.5) | 15 (39.5) | 67 (57.8) | 29 (63.0) |  | 29 (56.9) | 34 (64.2) | 25 (54.4) | 23 (46.0) |  |
| Chest computed tomography |  |  |  |  |  |  |  |  |  |  |
| No ground glass opacities | 8/17 (47.1) | 2/4 (50.0) | 5/9 (55.6) | 1/4 (25.0) | 0.376 | 1/1 (100) | 2/7 (28.6) | 3/6 (50.0) | 2/3 (66.7) | 0.806 |
| Unilateral ground glass opacities | 3/17 (17.7) | 0/4 (0.0) | 1/9 (11.1) | 2/4 (50.0) |  | 0/1 (0.0) | 2/7 (28.6) | 1/6 (16.7) | 0/3 (0.0) |  |
| Bilateral ground glass opacities | 6/17 (35.3) | 2/4 (50.0) | 3/9 (33.3) | 1/4 (25.0) |  | 0/1 (0.0) | 3/7 (42.8) | 2/6 (33.3) | 1/3 (33.3) |  |

Notes: (1) The laboratory values and imaging findings are presented as no./total no. (%) unless specified differently. (2) p-values refer to Chi-square test/ANOVA and the letters denote the columns where a statistically significant pairwise comparison exists (Bonferroni's method)
Abbreviations: BMI=body mass index, IQR=interquartile range, no.=number, g=gram, ng=nanogram, ug=microgram, mg=milligram, L=liter, uL=microliter, dL=deciliter, mL=milliliter, U=unit, AST=aspartate aminotransferase, ALT=alanine aminotransferase, CK=creatinine kinase, LDH=lactic dehydrogenase

In total, 24% of our cohort died during hospitalization, with higher rates among individuals with severe obesity (BMI<25 kg/m^2^: 31.6%, BMI 25-34 kg/m^2^: 17.2%, BMI≥35 kg/m^2^: 34.8%, p= 0.030). Similarly, patients with severe obesity were more likely to undergo intubation (BMI<25 kg/m^2^: 18.4%, BMI 25-34 kg/m^2^: 16.4%, BMI≥35 kg/m^2^: 34.8%, p= 0.032). In total, 45% of our patients had increasing oxygen requirements during hospital stay without significant differences among BMI groups. Twenty-two percent developed ARDS and 16% spent at least one night in the ICU. In-hospital outcomes are presented in **Table 4**.

**Table 4.** In-hospital outcomes

| Outcomes | All patients | BMI group |  |  |  | Age group |  |  |  |  |
| --- | --- | --- | --- | --- | --- | --- | --- | --- | --- | --- |
|  |  | BMI<25<br>(N=38) | BMI 25-34<br>(N=116) | BMI≥35<br>(N=46) | p-value | ≤50<br>(N=51) | 51-64<br>(N=53) | 65-73<br>(N=46) | ≥74<br>(N=50) | p-value |
| no./total no. (%) | N=200 |  |  |  |  |  |  |  |  |  |
| Mortality | 48 (24.0) | 12 (31.6) | 20 (17.2)* | 16(34.8)* | 0.030 | 6 (11.8)* | 12 (22.6) | 10 (21.7) | 20 (40.0)* | 0.010 |
| Intubation | 42 (21.0) | 7 (18.4) | 19 (16.4)* | 16 (34.8)* | 0.032 | 5 (9.8)* | 12 (22.6) | 15 (32.6)* | 10 (20.0) | 0.052 |
| ↑ O<sub>2</sub> requirement | 90 (45.0) | 17 (44.7) | 46 (39.7) | 27 (58.7) | 0.090 | 18 (35.3) | 25 (47.2) | 22 (47.8) | 25 (50.0) | 0.441 |
| ARDS | 45 (22.5) | 6 (15.8) | 25 (21.6) | 14 (30.4) | 0.259 | 9 (17.7) | 10 (18.9) | 13 (28.3) | 13 (26.0) | 0.509 |
| ICU | 32 (16.0) | 3 (7.89) | 18 (15.5) | 11 (23.9) | 0.134 | 5 (9.8) | 11 (20.8) | 12 (26.1)* | 4 (8.0)* | 0.042 |
| AKI | 70 (35.0) | 13 (34.2) | 39 (33.6) | 18 (39.4) | 0.798 | 7 (13.7)* | 19 (35.9) | 20 (43.5) | 24 (48.0)* | 0.002 |
| RRT | 16 (8.0) | 1 (2.6) | 9 (7.8) | 6 (13) | 0.216 | 2 (3.9) | 5 (9.6) | 5 (10.9) | 4 (8.00) | 0.606 |
| Length of stay median (IQR) - days | 6 (4-10) | 6 (4-12) | 5 (4-10) | 6 (4-9) | 0.924 | 5 (4-7) | 7 (5-12) | 6 (4-11.5) | 6 (4-11) | 0.273 |

Notes: (1) The outcomes are presented as no. (%) unless specified differently. (2) p-values refer to Chi-square test/ANOVA and stars denote the number of statistically significant pairwise comparisons using Bonferroni's method
Abbreviations and symbols: BMI=body mass index, IQR=interquartile range, no.=number, O<sub>2</sub>=oxygen, ↑=increasing, ARDS=acute respiratory distress syndrome, ICU=intensive care unit, AKI=acute kidney injury, RRT=renal replacement therapy

### Logistic Regression Analyses

#### In-hospital Mortality

The univariate associations with in-hospital mortality were examined for all the baseline demographic and clinical characteristics. Increasing age (analyzed in quartiles), male sex, BMI≥35 kg/m^2^ (reference: BMI 25-34 kg/m^2^), heart failure, CAD, and CKD or ESRD were found to have a significant univariate association (**Table 5**). The following variables were not shown to be statistically significant in the univariate associations: hypertension, hyperlipidemia, obstructive sleep apnea, diabetes, and smoking (**Table 5**). In the multivariable analysis (model 3), male sex (OR: 2.74; 95% CI: 1.25-5.98; p=0.011), increasing age (OR: 1.73; 95% CI: 1.13 - 2.63; p=0.011), and BMI≥35 kg/m^2^ (OR: 3.78; 95% CI: 1.45 - 9.83; p=0.006) were found to have significant associations.

**Table 5.** Univariate and multivariate logistic regression analyses for in-hospital mortality

| Variable | Univariate | Multivariate |  |  |
| --- | --- | --- | --- | --- |
|  | OR, 95% CI, p value | (1)<br>OR, 95% CI, p value | (2)<br>OR, 95% CI, p value | (3)<br>OR, 95% CI, p value |
| Male sex | 2.31 (1.18 - 4.54) p=0.015 |  | 2.76 (1.29 - 5.93) p=0.009 | 2.74 (1.25 - 5.98) p=0.011 |
| Age (quartiles) | 1.61 (1.19 - 2.20) p=0.002 | 1.75 (1.23 - 2.49) p=0.002 | 1.74 (1.15 - 2.65) p=0.009 | 1.73 (1.13 - 2.63) p=0.011 |
| African American or Hispanic | 0.45 (0.20 - 1.04) p=0.062 |  |  |  |
| BMI (<25) (25-34) (≥35) | 1.15 (0.65 - 2.02) p=0.637 |  |  |  |
| BMI (<25) (25-29) (≥30) | 0.79 (0.52 - 1.21) p=0.281 |  |  |  |
| BMI (Ref. 25-34) <25 | 2.22 (0.96 - 5.13) p=0.063 | 1.57 (0.68 - 3.66) p=0.294 | 1.31 (0.50 - 3.45) p=0.587 | 1.37 (0.52 - 3.64) p=0.527 |
| ≥35 | 2.56 (1.18 - 5.57) p=0.018 | 3.35 (1.43 - 7.87) p=0.005 | 3.94 (1.56 - 9.92) p=0.004 | 3.78 (1.45 - 9.83) p=0.006 |
| Heart failure | 3.18 (1.46 - 6.93) p=0.004 |  | 1.46 (0.52 - 4.13) p=0.471 | 1.43 (0.50 - 4.06) p=0.501 |
| Coronary artery disease | 2.88 (1.31 - 6.34) p=0.008 |  | 1.56 (0.57 - 4.30) p=0.389 | 1.53 (0.54 - 4.34) p=0.421 |
| Diabetes | 1.76 (0.91 - 3.40) p=0.091 |  |  | 1.16 (0.55 - 2.44) p=0.698 |
| CKD or ESRD | 2.14 (1.08 - 4.24) p=0.029 |  | 1.16 (0.50 - 2.69) p=0.723 | 1.15 (0.49 - 2.68) p=0.746 |
| COPD | 3.39 (1.48 - 7.80) p=0.004 |  | 1.85 (0.75 - 4.56) p=0.182 | 2.05 (0.76 - 5.51) p=0.156 |
| Residence status (community vs SNF) | 0.90 (0.42 - 1.91) p=0.779 |  |  |  |
| Current or former smoker | 1.19 (0.60 - 2.36) p=0.622 |  |  | 0.83 (0.37 - 1.87) p=0.647 |
| Alcohol use | 0.61 (0.17 - 2.21) p=0.450 |  |  |  |
| Intravenous drug use | 0.44 (0.05 - 3.69) p=0.450 |  |  |  |
| ACEI or ARB use prior to admission | 0.78 (0.38 - 1.61) p=0.503 |  |  |  |
| Cerebrovascular disease | 0.92 (0.32 - 2.66) p=0.883 |  |  |  |
| Hypertension | 0.93 (0.44 - 1.98) p=0.853 |  |  |  |
| Hyperlipidemia | 1.09 (0.57 - 2.10) p=0.789 |  |  |  |
| Asthma | 0.51 (0.17 - 1.56) p=0.238 |  |  |  |
| Obstructive sleep apnea | 2.27 (0.76 - 6.76) p=0.141 |  |  |  |
| Active malignancy | 1.88 (0.53 - 6.75) p=0.331 |  |  |  |
| On immunosuppressive therapy | 0.66 (0.18 - 2.40) p=0.525 |  |  |  |
| Any Disorder | 2.17 (0.85 - 5.54) p=0.104 |  |  |  |
| Income (above sample median) | 1.56 (0.80 - 3.00) p=0.188 |  |  |  |

Notes: (1) Multivariate analysis with age (quartiles), BMI <25 and BMI ≥35 as regressors, (2) Multivariate analysis with addition of all statistically significant variables of the univariate analysis as regressors, (3) Multivariate Analysis with addition of smoking and diabetes clinical important ones as regressors. Median income was estimated by zip codes. Age in years. BMI in kg/m<sup>2</sup>. For all calculations heteroscedastic adjusted standard errors were used.
Abbreviations: BMI=body mass index, CKD=chronic kidney disease, ESRD=end-stage renal disease, COPD=chronic obstructive pulmonary disease, SNF=skilled nursing facility, ACEi=angiotensin-converting-enzyme inhibitor, ARB=angiotensin II receptor blocker, CI=confidence interval, Ref.=reference

#### Increasing Oxygen Requirements

Male sex, current or former smoking and BMI≥35 kg/m^2^ (reference: BMI 25-34 kg/m^2^) were found to have a significant univariate association with increasing oxygen requirements (**Table 6**). In the multivariate analysis (model 3), male sex (OR: 2.77; 95% CI: 1.48 - 5.19; p=0.001), increasing age analyzed in quartiles (OR: 1.38; 95% CI: 1.01 - 1.89; p=0.042), BMI > 35 kg/m^2^ (OR: 3.09; 95% CI: 1.43 - 6.69; p=0.004), and current or former smoking (OR: 2.10; 95% CI: 1.07 - 4.10; p=0.031) were significant predictors (**Table 6**).

**Table 6.** Univariate and multivariate logistic regression analyses for increasing oxygen requirements

| Variable | Univariate | Multivariate |  |  |
| --- | --- | --- | --- | --- |
|  | OR, 95% CI, p value | (1)<br>OR, 95% CI, p value | (2)<br>OR, 95% CI, p value | (3)<br>OR, 95% CI, p value |
| Male sex | 2.25 (1.27 - 3.98) p=0.005 |  | 2.71 (1.48 - 4.98) p=0.001 | 2.77 (1.48 - 5.19) p=0.001 |
| Age (quartiles) | 1.20 (0.93 - 1.54) p=0.154 | 1.28 (0.98 - 1.67) p=0.070 | 1.37 (1.03 - 1.82) p=0.033 | 1.38 (1.01 - 1.89) p=0.042 |
| African American or Hispanic | 0.86 (0.39 - 1.89) p=0.702 |  |  |  |
| BMI (<25) (25-34) (≥35) | 1.37 (0.88 - 2.13) p=0.170 |  |  |  |
| BMI (<25) (25-29) (≥30) | 1.09 (0.76 - 1.58) p=0.628 |  |  |  |
| BMI (Ref. 25-34) <25 | 1.23 (0.59 - 2.59) p=0.582 | 1.04 (0.49 - 2.20) p=0.928 | 0.83 (0.38 - 1.80) p=0.635 | 0.95 (0.43 - 2.11) p=0.893 |
| ≥35 | 2.16 (1.08 - 4.34) p=0.030 | 2.37 (1.17 - 4.82) p=0.017 | 2.99 (1.44 - 6.21) p=0.003 | 3.09 (1.43 - 6.69) p=0.004 |
| Heart failure | 0.96 (0.45 - 2.02) p=0.910 |  |  | 0.56 (0.22 - 1.44) p=0.229 |
| Coronary artery disease | 1.37 (0.65 - 2.90) p=0.413 |  |  | 1.23 (0.51 - 2.94) p=0.641 |
| Diabetes | 1.46 (0.82 - 2.58) p=0.198 |  |  | 1.13 (0.60 - 2.14) p=0.710 |
| CKD or ESRD | 1.46 (0.79 - 2.71) p=0.224 |  |  | 1.00 (0.48 - 2.12) p=0.991 |
| COPD | 1.77 (0.79 - 3.97) p=0.168 |  |  | 1.02 (0.40 - 2.59) p=0.964 |
| Residence status (community vs SNF) | 1.14 (0.59 - 2.21) p=0.701 |  |  |  |
| Current or former smoker | 1.86 (1.02 - 3.39) p=0.042 |  | 2.10 (1.11 - 3.96) p=0.022 | 2.10 (1.07 - 4.10) p=0.031 |
| Alcohol use | 2.05 (0.76 - 5.54) p=0.157 |  |  |  |
| Intravenous drug use | 1.23 (0.30 - 5.09) p=0.773 |  |  |  |
| ACEI or ARB use prior to admission | 0.84 (0.46 - 1.53) p=0.560 |  |  |  |
| Cerebrovascular disease | 0.83 (0.34 - 2.04) p=0.684 |  |  |  |
| Hypertension | 0.86 (0.45 - 1.64) p=0.642 |  |  |  |
| Hyperlipidemia | 1.22 (0.69 - 2.13) p=0.496 |  |  |  |
| Asthma | 0.57 (0.24 - 1.34) p=0.195 |  |  |  |
| Obstructive sleep apnea | 1.08 (0.37 - 3.10) p=0.893 |  |  |  |
| Active malignancy | 1.50 (0.44 - 5.10) p=0.516 |  |  |  |
| On immunosuppressive therapy | 0.84 (0.31 - 2.32) p=0.741 |  |  |  |
| Any Disorder | 1.63 (0.80 - 3.29) p=0.177 |  |  |  |
| Income (above sample median) | 1.00 (0.57 - 1.75) p=1.000 |  |  |  |

Notes: (1) Multivariate analysis with age (quartiles), BMI <25 and BMI ≥35 as regressors, (2) Multivariate analysis with addition of all statistically significant variables of the univariate analysis as regressors, (3) Multivariate Analysis with addition of smoking and diabetes clinical important ones as regressors. Median income was estimated by zip codes. Age in years. BMI in kg/m<sup>2</sup>. For all calculations heteroscedastic adjusted standard errors were used.
Abbreviations: BMI=body mass index, CKD=chronic kidney disease, ESRD=end-stage renal disease, COPD=chronic obstructive pulmonary disease, SNF=skilled nursing facility, ACEi=angiotensin-converting-enzyme inhibitor, ARB=angiotensin II receptor blocker, CI=confidence interval, Ref.=reference

#### Intubation

Male sex, and BMI≥35 kg/m^2^ (reference: BMI 25-34.9 kg/m^2^) were found to have a significant univariate association with intubation (**Table 7**). In the multivariate analysis (model 3), male sex (OR: 3.35; 95% CI: 1.51 - 7.46, p=0.003), increasing age analyzed in quartiles (OR: 1.50; 95% CI:1.05 - 2.12; p=0.025), and BMI≥35 kg/m^2^ (OR: 3.87; 95% CI: 1.47 - 10.18; p=0.006) were significant predictors (**Table 7**).

**Table 7.** Univariate and multivariate logistic regression analyses for intubation

| Variable | Univariate | Multivariate |  |  |
| --- | --- | --- | --- | --- |
|  | OR, 95% CI, p value | (1)<br>OR, 95% CI, p value | (2)<br>OR, 95% CI, p value | (3)<br>OR, 95% CI, p value |
| Male sex | 2.51 (1.23 - 5.14) p=0.012 |  | 3.39 (1.56 - 7.39) p=0.002 | 3.35 (1.51 - 7.46) p=0.003 |
| Age (quartiles) | 1.27 (0.97 - 1.68) p=0.087 | 1.45 (1.06 - 1.97) p=0.019 | 1.60 (1.15 - 2.22) p=0.005 | 1.50 (1.05 - 2.12) p=0.025 |
| African American or Hispanic | 0.65 (0.27 - 1.60) p=0.350 |  |  |  |
| BMI (<25) (25-34) (≥35) | 1.72 (0.94 - 3.12) p=0.077 |  |  |  |
| BMI (<25) (25-29) (≥30) | 0.79 (0.52 - 1.20) p=0.281 |  |  |  |
| BMI (Ref. 25-34) <25 | 1.15 (0.44 - 3.01) p=0.771 | 0.90 (0.33 - 2.44) p=0.829 | 0.79 (0.28 - 2.17) p=0.643 | 0.76 (0.26 - 2.22) p=0.613 |
| ≥35 | 2.72 (1.24 - 5.96) p=0.012 | 3.19 (1.42 - 7.17) p=0.005 | 4.06 (1.72 - 9.57) p=0.001 | 3.87 (1.47 - 10.18) p=0.006 |
| Heart failure | 1.45 (0.62 - 3.41) p=0.393 |  |  |  |
| Coronary artery disease | 1.25 (0.52 - 3.03) p=0.618 |  |  |  |
| Diabetes | 1.95 (0.98 - 3.88) p=0.058 |  |  | 1.26 (0.58 - 2.73) p=0.557 |
| CKD or ESRD | 1.30 (0.62 - 2.69) p=0.488 |  |  |  |
| COPD | 2.00 (0.83 - 4.82) p=0.125 |  |  |  |
| Residence status (community vs SNF) | 1.39 (0.59 - 3.27) p=0.446 |  |  |  |
| Current or former smoker | 1.56 (0.77 - 3.15) p=0.217 |  |  | 1.66 (0.76 - 3.62) p=0.206 |
| Alcohol use | 1.51 (0.50 - 4.51) p=0.463 |  |  |  |
| Intravenous drug use | 1.27 (0.25 - 6.54) p=0.778 |  |  |  |
| ACEI or ARB use prior to admission | 0.64 (0.29 - 1.40) p=0.261 |  |  |  |
| Cerebrovascular disease | 0.56 (0.16 - 2.01) p=0.376 |  |  |  |
| Hypertension | 0.63 (0.30 - 1.35) p=0.239 |  |  |  |
| Hyperlipidemia | 1.98 (0.99 - 3.96) p=0.055 |  |  | 1.66 (0.78 - 3.55) p=0.188 |
| Asthma | 0.62 (0.20 - 1.90) p=0.401 |  |  |  |
| Obstructive sleep apnea | 2.76 (0.92 - 8.27) p=0.070 |  |  | 1.15 (0.40 - 3.35) p=0.791 |
| Active malignancy | 1.44 (0.36 - 5.71) p=0.602 |  |  |  |
| On immunosuppressive therapy | 0.79 (0.22 - 2.90) p=0.724 |  |  |  |
| Any Disorder | 2.26 (0.83 - 6.19) p=0.112 |  |  |  |
| Income (above sample median) | 1.27 (0.64 - 2.53) p=0.489 |  |  |  |

Notes: (1) Multivariate analysis with age (quartiles), BMI <25 and BMI ≥35 as regressors, (2) Multivariate analysis with addition of all statistically significant variables of the univariate analysis as regressors, (3) Multivariate Analysis with addition of smoking and diabetes clinical important ones as regressors. Median income was estimated by zip codes. Age in years. BMI in kg/m<sup>2</sup>. For all calculations heteroscedastic adjusted standard errors were used.
Abbreviations: BMI=body mass index, CKD=chronic kidney disease, ESRD=end-stage renal disease, COPD=chronic obstructive pulmonary disease, SNF=skilled nursing facility, ACEi=angiotensin-converting-enzyme inhibitor, ARB=angiotensin II receptor blocker, CI=confidence interval, Ref.=reference

### Interaction Analysis

Given the significant associations that we noticed for male sex, age and BMI≥35 kg/m^2^ with the outcomes that we examined, we performed an interaction analysis for this set of variables. Two different interactions were tested; sex with BMI and age with BMI. Both of them were not significant.

## Discussion

Our study described the baseline characteristics, clinical features, and early outcomes of the first 200 patients who were hospitalized due to COVID-19 in our institution that mainly serves African American and Hispanic population. This is the first study that has performed joint evaluation of age, gender, obesity and multiple comorbidities that have previously been linked with adverse outcomes. The main findings can be summarized as following: 1) the in-hospital mortality was 24% with only 3% patients still hospitalized on the 21-day follow-up 2) severe obesity (BMI≥35 kg/m^2^), increasing age, and male sex have an independent association with mortality and need for intubation 3) severe obesity (BMI≥35 kg/m^2^), increasing age, male sex, and smoking have an independent association with increasing oxygen requirements during hospitalization.

Older age and male sex have already been described as risks factor for severe disease and death in patients with COVID-19 [5,11-16], although large outcome studies are needed to assess the latter. The most interesting finding of our analysis is that severe obesity is a significant factor for severe respiratory disease and death in hospitalized patients with COVID-19. It should be pointed out that this association remained significant after adjusting for several clinical entities, such as diabetes, coronary artery disease, heart failure, COPD, CKD or ESRD, and smoking, which indicates that obesity may predispose to negative outcomes independently. To the best of our knowledge, no other studies to date have shown an independent and direct association of severe obesity to mortality in patients with COVID-19; however, some preliminary published data have linked obesity to severe COVID-19. A large cohort from New York City depicted that obesity is strongly associated with progression to critical illness with substantially higher odds ratio than any cardiovascular or pulmonary disease (BMI 30-40 kg/m^2^ OR: 1.38; 95% CI: 1.03-1.85; BMI>40 kg/m^2^ OR: 1.73; 95% CI 1.03-2.90) [17]. A cohort from China revealed that obesity significantly increases the risk for developing severe pneumonia in the setting of COVID-19 (OR: 3.42; 95% CI: 1.42-8.27) [18]. Another report from China indicated that the presence of obesity in patients with metabolic-associated fatty liver disease is associated with an almost 6-fold increased risk of severe COVID-19 [19]. Obesity could partially explain why the mortality rate for COVID-19 is higher in countries with higher prevalence of obesity, such as Italy, as compared to China and Japan [20]. Variables such as diabetes and other cardiovascular and pulmonary comorbidities were found to have a significant association in prior reports from China and Italy [11-14]. However, most of these were obtained from univariate estimates only. In our multivariate analyses it was shown that these comorbidities are likely epiphenomena since they are not independent from male gender, older age or obesity which may be the underlying link.

Our findings on the association of severe obesity to mortality in COVID-19 are not unanticipated given our prior experience from the 2009 pandemic influenza (H1N1) disease. Morgan et al. reported significant association of obesity to death in adult patients without recognized pre-existing medical conditions hospitalized with H1N1 influenza disease (obesity OR: 3.1; 95% CI: 1.5 - 6.6); morbid obesity OR: 7.6; 95% CI: 2.1 - 27.9) [21]. Obesity leads to increased work of breathing by augmenting the airway resistance and is associated with decreased expiratory reserve volume, functional capacity, and pulmonary compliance [22,23]. Central obesity results in decreased diaphragmatic excursion in supine patients compromising ventilation [23]. Moreover, obesity is a chronic inflammatory state with increased circulating levels of pro-inflammatory cytokines, including interleukin-6, and is known to impair the immune system [24, 25]. The observed association between severe obesity and mortality may also underlie the observed association between low vitamin D levels, which are low in the obese, and mortality and needs to be explored further [25].

The in-hospital mortality rate on 21-day follow-up in our cohort is 24%. Three inpatient cohorts from China reported in-hospital mortality rates of 28.2%, 11.7%, and 17%, respectively [5,11,12]. A large cross-sectional study from New York City, which did not include hospitals located in the Bronx, reported a 14.6% mortality rate in the inpatient sample to the time of the analysis, however 35.9% of the patients were still hospitalized, which indicates that the final inpatient mortality rate may be actually higher [16]. Large retrospective cohorts that will probably be published in the following months will accurately estimate the in-hospital mortality in the Bronx and elsewhere.

In total, 33.6% of adults in the Bronx are obese, a number which is much higher than all other NYC boroughs, while the prevalence of obesity nationally is 20% [9,10]. Additionally, the prevalence of obesity is higher among individuals of lower socioeconomic status (>35%) and substantially higher in non-Hispanic blacks (36%) and Hispanic (35.4%) as compared to non-Hispanic whites (19.1%) [9]. The patient population that our institution serves mainly includes patients of lower socioeconomic status and people that identify themselves as African American or Hispanic. The combination of these two made up 85.5% of our current study population. The median income, as estimated by the zip codes based on publicly available data provided by the internal revenue service, was not found to be a risk factor for worse outcomes in our study, which is explained by the fact that the Bronx is a homogenous area of low income.

We report that active or prior smoking was associated with increasing oxygen requirements during hospitalization. This is in contrast to the results of a recent meta-analysis of five studies from China which concluded that active smoking did not seem to be significantly associated with higher risk of progressing to severe COVID-19 [26]. Large observational studies and future meta-analyses will elucidate the association of smoking with COVID-19 severity and other outcomes.

To the best of our knowledge, this is the first study to date showing the independent association of severe obesity to in-hospital mortality in patients with COVID-19. One of the strengths of the current study is that our included patients represent underserved and economically disadvantaged minorities; thus, revealing the early outcomes of COVID-19 in this vulnerable population and usually underreported and underrepresented in clinical research. Additionally, two researchers independently and blindly collected data which reduces errors and bias. On the other hand, our study has several limitations. First, our sample was relatively small, but given the nature of the evolving pandemic, it was of paramount importance to make our early data and findings widely available as soon as possible, especially given the lack of data up to date in COVID-19 in minorities and underserved population. Second, this was a real-world study with a retrospective design utilizing the electronic medical records, which is suboptimal compared to a prospective study that could have more accurate follow-up assessment. Third, the rapidly changing management of COVID-19 might have affected our results but it highly unlikely that could have differentials affected associations between obesity and mortality. Fourth, we handled BMI as a categorical variable in the regression analysis. This can lead to suboptimal conclusions, but we think that specific cut-offs, following established clinical guidelines on obesity, may be of more interest and ease for the clinicians compared to interpretation of continuous variables in a regression model.

In conclusion in this early cohort of hospitalized patients with COVID-19 in an underserved, minority-predominant population in the Bronx, we found that severe obesity was associated with higher in-hospital mortality even after adjusting for other pertinent potential confounding factors. Particular attention should be paid in protection of this population given the higher chance for negative outcomes once they are diagnosed with the disease. In addition, obese patients diagnosed with COVID-19 should be treated with extra caution given the possible higher risk for adverse outcomes. While we recognize the limitations, we hope that our study will stimulate additional researchers to further study the effect of obesity in COVID-19 and outcomes of minorities diagnosed with COVID-19. Larger cohort studies are needed to confirm our data and pilot clinical trials are needed to assess whether pharmacotherapy for obesity and its comorbidities may improve outcomes in the short or the long term.

**Figure 1:**
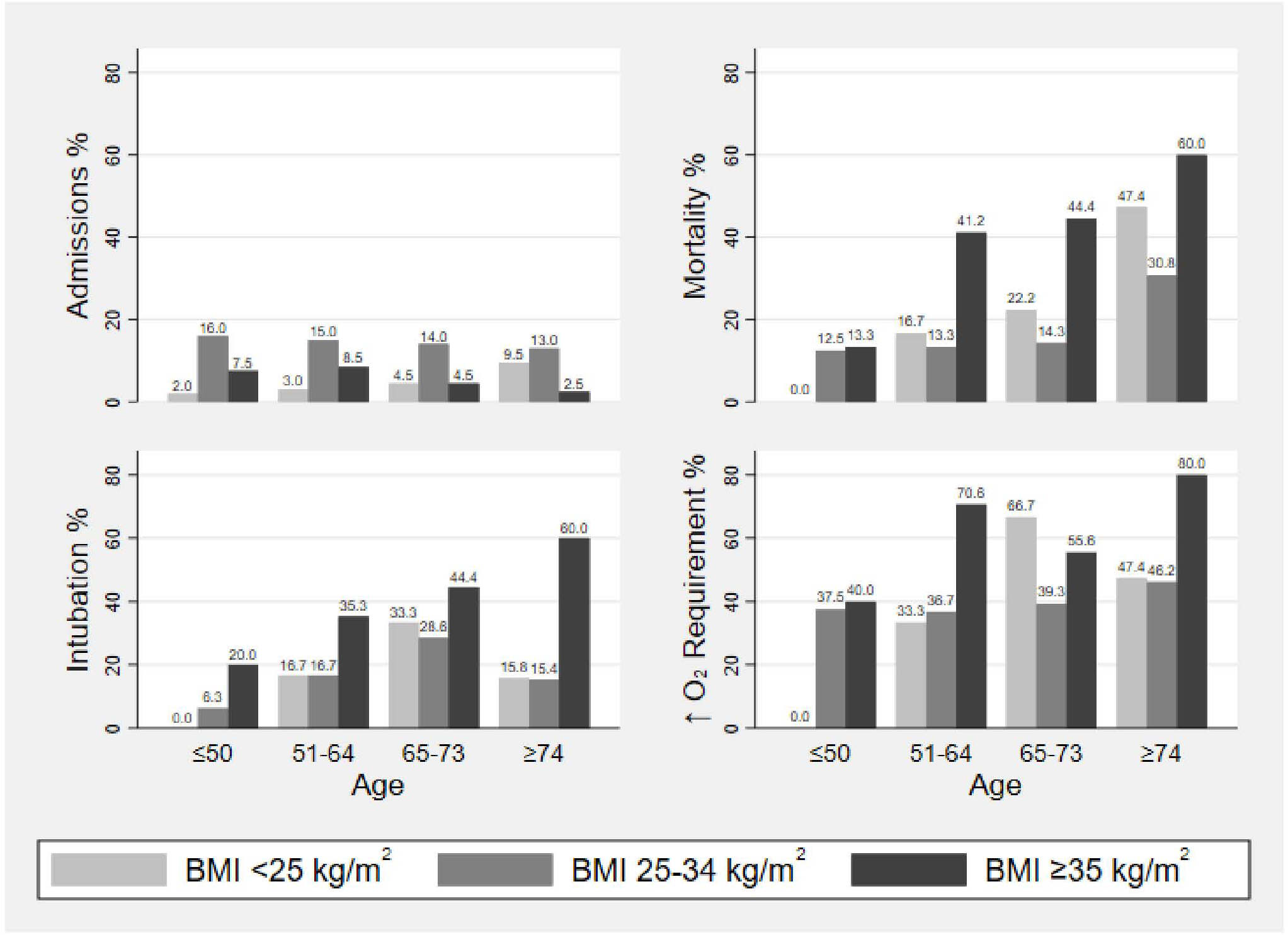
Study population, in-hospital mortality and secondary outcomes per age group (≤50, 51-64, 65-73, and ≥ 74 years) and body mass index (BMI <25, 25-34, and ≥ 35 kg/m^2^)

## Data Availability

Dataset is available upon request at the first author

## Abbreviations

COVID-19: coronavirus disease 2019
SARS-CoV-2: acute respiratory syndrome coronavirus 2
BMI: body mass index
OR: odds ratio
CI: confidence interval
EMR: electronic medical record
SNF: skilled nursing facility

## Authors contributions

Study concept and design: Leonidas Palaiodimos, Damianos G. Kokkinidis, Weijia Li and Christos S. Mantzoros

Acquisition of data: Leonidas Palaiodimos, Weijia Li, and Jennifer Ognibene

Analysis and interpretation of data: Leonidas Palaiodimos, Damianos G. Kokkinidis, Dimitrios Karamanis, Jennifer Ognibene, Shitij Arora, William N Southern, and Christos S. Mantzoros

Drafting of the manuscript: Leonidas Palaiodimos and Damianos G. Kokkinidis

Critical revision of the manuscript for important intellectual content: Shitij Arora, William N. Southern, and Christos S. Mantzoros

Study Supervision: Christos S. Mantzoros

All authors contributed to the manuscript for important intellectual contents and approved the submission.

## References

[1]. World Health Organization. Coronavirus disease 2019 (COVID-19) situation report – 87. Updated April 27, 2020. Accessed April 27, 2020. https://www.who.int/docs/default-source/coronaviruse/situation-reports/20200427-sitrep-98-covid-19.pdf?sfvrsn=90323472_4

[2]. Bhatraju PK, Ghassemieh BJ, Nichols M, Kim R, Jerome KR, Nalla AK, Greninger AL, Pipavath S, Wurfel MM, Evans L, Kritek PM. Covid-19 in critically ill patients in the Seattle region—case series. New England Journal of Medicine. 2020 Mar 30.

[3]. Centers for Disease Control and Prevention. Coronavirus disease 2019 (COVID-19). Cases in the US. Updated April 26, 2020. Accessed April 27, 2020. https://www.cdc.gov/coronavirus/2019-ncov/cases-updates/cases-in-us.html

[4]. New York City Department of Health. Coronavirus disease 2019 (COVID-19). COVID-19: Data. Updated April 27, 2020. Accessed April 27, 2020. https://www1.nyc.gov/site/doh/covid/covid-19-data.page

[5]. Zhou F, Yu T, Du R, Fan G, Liu Y, Liu Z, Xiang J, Wang Y, Song B, Gu X, Guan L. Clinical course and risk factors for mortality of adult inpatients with COVID-19 in Wuhan, China: a retrospective cohort study. The Lancet. 2020 Mar 11

[6]. Grasselli G, Zangrillo A, Zanella A, Antonelli M, Cabrini L, Castelli A, Cereda D, Coluccello A, Foti G, Fumagalli R, Iotti G. Baseline characteristics and outcomes of 1591 patients infected with SARS-CoV-2 admitted to ICUs of the Lombardy region, Italy. Jama. 2020 Apr 6.

[7]. Yancy CW. COVID-19 and African Americans. Jama. doi:10.1001/jama.2020.6548

[8]. County Health Rankings and Roadmaps. 2019 County Health Rankings Key Findings Report. Published March 2019. Accessed April 16, 2020. https://www.countyhealthrankings.org/reports/2019-county-health-rankings-key-findings-report

[9]. Montefiore’s Office of Community & Population Health Bronx Community Health Dashboard: Nutrition, Physical Activity and Obesity. Updated January 16, 2018. Accessed April 16, 2020. https://www.montefiore.org/documents/communityservices/OCPH-Dashboard-Obesity.pdf

[10]. Centers for Disease Control and Prevention. Adult Obesity Prevalence Maps. Division of Nutrition, Physical Activity, and Obesity, National Center for Chronic Disease Prevention and Health Promotion. Updated August, 7 2019. Accessed April 16, 2020. https://www.cdc.gov/obesity/data/index.html.

[11]. Du RH, Liang LR, Yang CQ, Wang W, Cao TZ, Li M, Guo GY, Du J, Zheng CL, Zhu Q, Hu M. Predictors of Mortality for Patients with COVID-19 Pneumonia Caused by SARS-CoV- 2: A Prospective Cohort Study. European Respiratory Journal. 2020 Jan 1.

[12]. Fu L, Fei J, Xiang HX, Xiang Y, Tan ZX, Li MD, Liu FF, Liu HY, Zheng L, Li Y, Zhao H. Analysis of Death Risk Factors Among 200 COVID-19 Patients in Wuhan, China: A Hospital-Based Case-Cohort Study. Fang-Fang and Liu, Hong-Yan and Zheng, Ling and Li, Ying and Zhao, Hui and Xu, De-Xiang, Analysis of Death Risk Factors Among. 2020 Mar 6;200.

[13]. Onder G, Rezza G, Brusaferro S. Case-fatality rate and characteristics of patients dying in relation to COVID-19 in Italy. Jama. 2020 Mar 23.

[14]. Zhang J, Wang X, Jia X, Li J, Hu K, Chen G, Wei J, Gong Z, Zhou C, Yu H, Yu M. Risk factors for disease severity, unimprovement, and mortality of COVID-19 patients in Wuhan, China. Clinical Microbiology and Infection. 2020 Apr 15.

[15]. Mo P, Xing Y, Xiao Y, Deng L, Zhao Q, Wang H, Xiong Y, Cheng Z, Gao S, Liang K, Luo M. Clinical characteristics of refractory COVID-19 pneumonia in Wuhan, China. Clinical Infectious Diseases. 2020 Mar 16.

[16]. Onder G, Rezza G, Brusaferro S. Case-fatality rate and characteristics of patients dying in relation to COVID-19 in Italy. Jama. 2020 Mar 23.

[17]. Petrilli CM, Jones SA, Yang J, Rajagopalan H, O’Donnell L, Chernyak Y, Tobin KA, Cerfolio RJ, Francois F, Horwitz LI. Factors associated with hospitalization and critical illness among 4,103 patients with Covid-19 disease in New York City. Preprint doi: 10.1101/2020.04.08.20057794

[18]. Cai Q, Chen F, Luo F, Liu X, Wang T, Wu Q, He Q, Wang Z, Liu Y, Chen J, Liu L, Xu L. Obesity and COVID-19 Severity in a Designated Hospital in Shenzhen, China. Preprint doi: 10.2139/ssrn.3556658

[19]. Zheng KI, Gao F, Wang XB, Sun QF, Pan KH, Wang TY, Ma HL, Liu WY, George J, Zheng MH. Obesity as a risk factor for greater severity of COVID-19 in patients with metabolic associated fatty liver disease. Metabolism. 2020 Apr 19: 154244.

[20]. Rebelos E, Moriconi D, Virdis A, Taddei S, Foschi D, Nannipieri M. Importance of metabolic health in the era of COVID-19. Metabolism-Clinical and Experimental. 2020 Apr 22.

[21]. Morgan OW, Bramley A, Fowlkes A, Freedman DS, Taylor TH, Gargiullo P, Belay B, Jain S, Cox C, Kamimoto L, Fiore A. Morbid obesity as a risk factor for hospitalization and death due to 2009 pandemic influenza A (H1N1) disease. PloS one. 2010;5(3).

[22]. Falagas ME, Kompoti M. Obesity and infection. The Lancet infectious diseases. 2006 Jul 1;6(7):438–46.

[23]. Dietz W, Santos-Burgoa C. Obesity and its Implications for COVID-19 Mortality. Obesity. 2020 Apr 1.

[24]. de Heredia FP, Gömez-Martínez S, Marcos A. Obesity, inflammation and the immune system. Proceedings of the Nutrition Society. 2012 May;71(2):332–8.

[25]. Muscogiuri G, Pugliese G, Barrea L, Savastano S, Colao A. Obesity: the "Achilles heel” for COVID-19? Metabolism-Clinical and Experimental. 2020 Apr 27.

[26]. Lippi G, Henry BM. Active smoking is not associated with severity of coronavirus disease 2019 (COVID-19). European journal of internal medicine. 2020 Mar 16.

